# Strengthening Policy Coding Methodologies to Improve COVID-19 Disease Modeling and Policy Responses: A Proposed Coding Framework and Recommendations

**DOI:** 10.1101/2020.08.12.20173740

**Authors:** Jeff Lane, Michelle M. Garrison, James Kelley, Priya Sarma, Aaron Katz

## Abstract

In recent months, multiple efforts have sought to characterize COVID-19 social distancing policy responses. These efforts have used various coding frameworks, but most have relied on binary coding that may not adequately describe the gradient in social distancing policies as states “re-open.” We developed a COVID-19 social distancing intensity framework that is sufficiently specific and sensitive to capture this gradient. Based on a review of policies from a 12-state sample, we developed a social distancing intensity framework consisting of 16 domains and intensity scales of 0-5 for each domain. We found that the states with the highest average daily intensity from our sample were Pennsylvania, Washington, Colorado, California, and New Jersey, with Georgia, Florida, Massachusetts, and Texas having the lowest. While some domains (such as restaurants and movie theaters) showed bimodal policy intensity distributions compatible with binary (yes/no) coding, others (such as childcare and religious gatherings) showed broader variability that would be missed without more granular coding. We also present a range of methodological recommendations to strengthen COVID-19 comparative policy coding efforts.

## Introduction

The first confirmed case of COVID-19 occurred in the United States in Washington State on January 20, 2020.(1) Non-pharmaceutical interventions, such as quarantines and mass social distancing, have been the primary public health strategy for blunting the initial wave of COVID-19. As confirmed case counts climbed, state, county, and municipal governments adopted policies recommending or requiring actions to reduce social density and slow the progression of the outbreak. The timing and intensity of social distancing policy responses has varied.

To date, multiple efforts have sought to rapidly code these social distancing policy responses for analysis. Social distancing policy coding has also been critical to COVID-19 disease models that have influenced policy decision-making whether to impose or ease social distancing approaches. These efforts have used a range of methodologies and frameworks to characterize and code these policy responses, resulting in a diversity of social distancing policy taxonomies and classification schemes. These efforts have characterized social distancing into taxonomies consisting of 1 domain (e.g., stay at home order in place) to upwards of six domains. Within domains, most methodologies used binary coding to state whether social distancing is in place for that domain or not. A small number of analyses have used an ordinal scale within domains to illustrate gradients of social distancing within a domain (e.g., different levels of restaurant restrictions). Exhibit 1 lists some of the social distancing policy coding efforts and describes the policy coding methodologies used.

**Exhibit 1:**
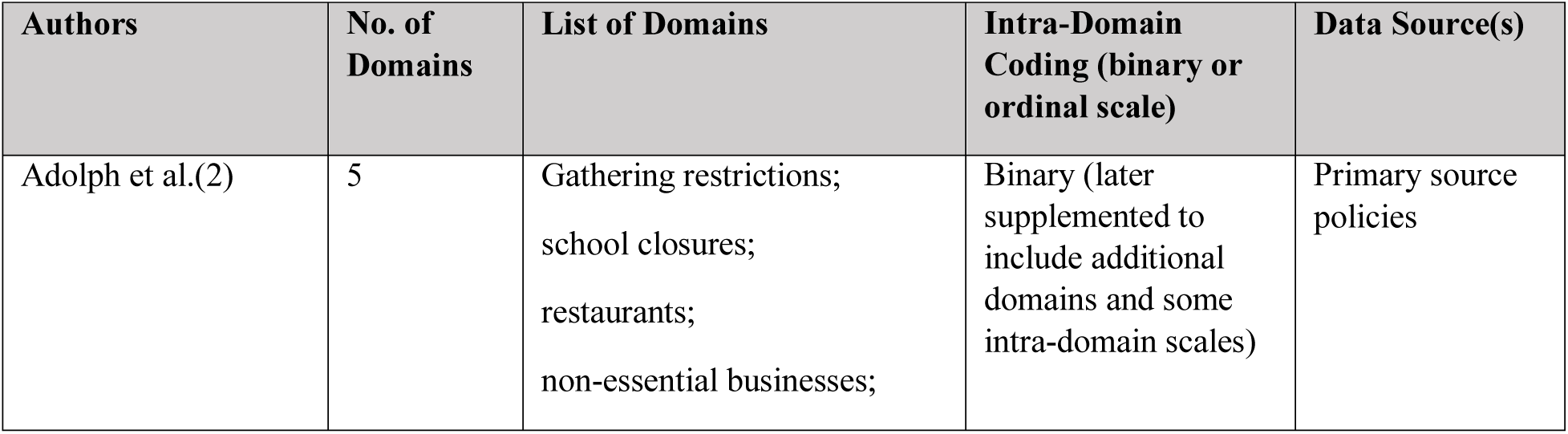
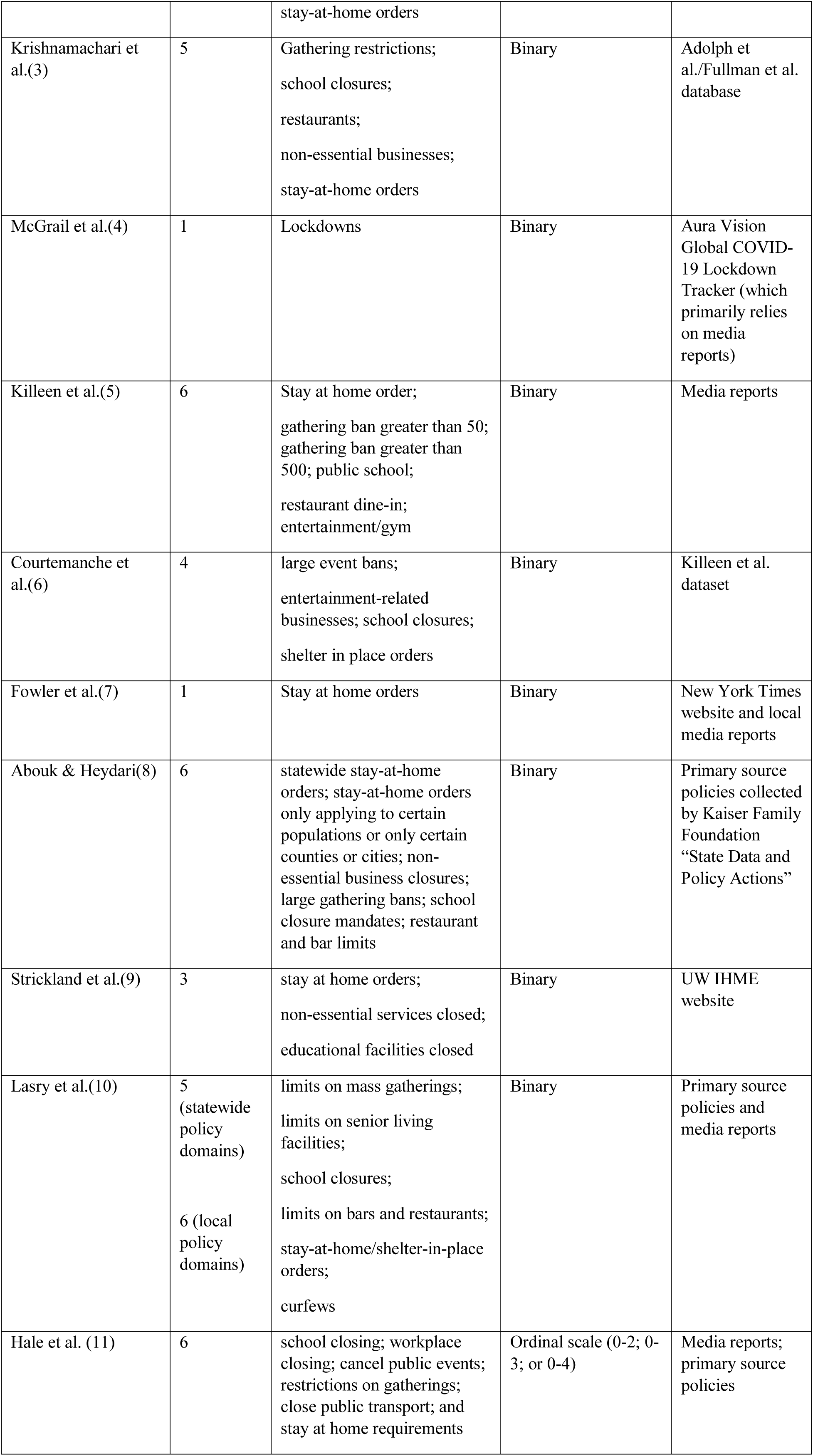
Existing COVID-19 Policy Responses Coding Frameworks

One of the first published analyses of social distancing policies was published on medRxiv by Adolph et al. on March 31, 2020.(2) The authors developed a COVID-19 policy database, publicly available on GitHub. The database was originally populated with data from the National Governor Association’s website and primary source policies. The authors used this database to “explore how political partisanship, COVID-19 caseload, and policy diffusion explain the timing of governors’ decisions to mandate social distancing.” This database initially tracked five social distancing policy responses: (1) recommendations or restrictions on gatherings, regardless of the size of gathering; (2) K-12 school closures; (3) restaurant restrictions on in-person dining; (4) non-essential business closures; and (5) mandatory stay-at-home orders. The GitHub site maintained by this group has been updated to include additional domains, including masking policies, and to use ordinal scales for some domains, such as gathering restrictions and masking policies.(12) Multiple subsequent analyses have since relied on this database. For example, Krishnamachari et al. used it to examine the effects of government implemented social distancing measures on the cumulative incidence rates of COVID-19 on the state level and in the 25 most populated cities.(3)

McGrail et al. used the Aura Vision Global COVID-19 Lockdown Tracker to quantify the spread rate of COVID-19 in 53 countries.(4) The Aura Vision Tracker characterizes social distancing policy responses as a single domain of “lockdown,” coded as the date the lockdown started and ending as of the date non-essential retail stores re-opened. The data sources cited were predominantly media reports.

Killeen et al. developed a dataset available on GitHub titled “County-level Socioeconomic Data for Predictive Modeling of Epidemiological Effects.”(13) This dataset is described as a “machine-readable dataset that aggregates relevant data from governmental, journalistic, and academic sources on the county level.” It used the following domains: stay at home order; gathering ban greater than 50; gathering ban greater than 500; public school closure; restaurant dine-in ban; and entertainment/gym closure.

Courtemanche et al. relied on the Killeen et al. dataset to evaluate the impact of social distancing on the growth rate of confirmed COVID-19 cases across US counties.(6) Their analysis examined four types of social distancing policies: large event bans, coded as the date of the first prohibition of at least 500 people; closure of entertainment-related businesses, coded as the date of the first closure of either restaurant dining areas (including bars) or gyms/entertainment centers; school closures; and shelter in place orders.

Fowler et al. relied on policy dates published on the New York Times website and local media reports to estimate the effect of a single domain (stay at home orders).(7) Abouk and Heydari measured the impact of six common policies on people’s presence at home and their mobility in different types of public places.(8) They used binary coding (in effect or not) in six domains: statewide stay-at-home orders; stay-at-home orders only applying to certain populations or only certain counties or cities; non-essential business closures; large gathering bans; school closures; and restaurant and bar service limits.

Strickland et al. assessed associations between state public health agency governance and timing and extent of implementing social distancing control measures during COVID-19 response.(9) They used binary coding for three domains: stay at home orders; non-essential services closed; and educational facilities closed. They merged these three domains into an aggregate “extensiveness of social distancing actions” score of 0-3 (0 = none; 1 = 1 measure; 2 = 2 measures; and 3 = 3 measures). This analysis relied on policy response data used by the University of Washington’s Institute for Health Metrics and Evaluation’s website.

Lasry et al. examined the timing of community mitigation and community mobility data in four cities: New York City, Seattle, San Francisco, and New Orleans.(10) They coded state and local policy responses in 6 domains: limits on mass gatherings; limits on senior living facilities, school closures, limits on bars and restaurants, stay-at-home/shelter-in-place orders, and curfews.

Hale et al. developed the Oxford COVID-19 Government Response Tracker that monitors 17 indicators of government responses to COVID-19 in 160 countries.(11) Six of these indicators focus on social distancing policies in the following domains: school closing; workplace closing; canceling public events; restricting gathering size; closing public transport; and stay at home requirements. They code domains using non-standardized ordinal scales of 0-2, 0-3, or 0-4, depending on the domain. They also use a binary flag to identify policies that have a targeted subnational geographic scope.

As of April 27, 2020, states have been lifting social distancing requirements. The “re-opening” process has often occurred in predefined stages leading to a gradient in social distancing that is hard to capture with a limited number of policy domains or a lack of a scale within domains. To help address such a fluid and complex policy environment, we sought to develop a COVID-19 policy coding framework that is sufficiently specific and sensitive to capture the gradient in social distancing policy responses. We also sought to identify methodological recommendations to strengthen the COVID-19 comparative policy analyses to better inform research, disease modeling, and policy decision-making. These recommendations may be useful for efforts to code other types of health policies to inform comparative analyses and modeling. We also hope that this more refined COVID-19 social distancing policy coding framework and our methodological recommendations for policy coding will lead to additional collaboration across COVID-19 policy tracking and analysis groups and be supplemented over time to include other key components of the COVID-19 policy response (e.g., mandated masking).

## Methods

We used a sample of the first twelve U.S. states to reach 100 confirmed COVID-19 cases (New York, Washington, California, New Jersey, Massachusetts, Louisiana, Florida, Colorado, Illinois, Georgia, Pennsylvania, Texas), which all of these states reached by March 17, 2020. The total population of these 12 states represented almost 55% of the entire population of the United States as of July 1, 2019.(14) Policies were identified by a search of public websites operated by state governments (e.g., governor or state department of health), and in some cases facilitated by reviewing the Kaiser Family Foundation list of state policy responses.

The inclusion criteria were: (1) directive issued by the Governor or state agency lead regarding COVID-19 documented in an Executive Order or state government website, including but not limited to Department of Health; (2) the primary purpose of the policy is to reduce social density with the goal of reducing community transmission of COVID-19; and (3) policies issued between March 11, 2020 and June 19, 2020. June 19 was selected as the end date, because that date marked 100 days after the World Health Organization declared COVID-19 a pandemic and approximately six months after the first confirmed COVID-19 case in the United States, which occurred on January 20, 2020. We excluded: (1) recommendations or directives issued orally only (e.g., at a press conference and not released as a press release on the governor’s website); (2) recommendations or directives issued by local governments (e.g., municipal or county government officials) or the U.S. federal government.

Primary source policies were reviewed longitudinally and characterized in an Excel sheet across a range of potential domains by two co-authors. One co-author (JL) coded all 12 states with other co-authors coding a subset of states (AK, JK, and PS). Differences in coding were discussed and resolved via consensus among the coders. Policy descriptors were added to the policy database to facilitate revisions to the coding framework based on the review of our 12-state sample. Policies were coded as of the effective date of the policy (not the announcement date). Policy descriptors were re-reviewed following finalization of the coding framework to ensure accuracy. Interpretation notes were added to the Excel datasheet to document key interpretations.

Location domains were identified inductively based on our review of social distancing policies in our sample states. Various domain definitions and scales were explored to develop a domain framework and intra-domain scale that would be sufficiently specific and sensitive to describe nuanced differences in policy responses. We subsequently reviewed two public COVID-19 policy tracking databases developed by Kaiser Family Foundation(15) and Raifman et al.(16) to validate our findings and identify any potential discrepancies.

We collected longitudinal, confirmed case count and mortality data from the Johns Hopkins University COVID Tracker database for descriptive statistics and data visualization. As the impact of policy on outbreak progression is not immediate, our data visualizations explored COVID-19 outcomes in relation to the concurrent policy intensity (i.e., the policy status on the same day as the outcomes) and the policy intensity as of 2 weeks prior to incidence rate outcomes and 6 weeks prior to mortality outcomes to better visualize an estimated lag between policy implementation and potential for impact. Infectious outcomes such as incidence and mortality rates can be presented as daily, weekly, monthly, or yearly rates. While daily rates can experience a lot of noise in day-to-day fluctuations – some of which may be driven in part by scheduling or other procedural issues – monthly rates may not be “real time” enough to pick up the impact of policy changes. To find a middle ground, here we present rolling 7-day averages – so the rate for each day is the incidence or mortality for the 7 days of which that date is the midpoint. While we would typically constrain all vertical axes to the same maxima if the goal was to directly compare incidence and mortality rates across states, here the goal is to examine the relationship between policy intensity and subsequent disease outcomes, and so we constrained the right-hand policy vertical axis to the maximum daily average policy intensity score but left the left-hand disease outcome vertical axis unconstrained.

## Results

Based on our analysis of the 12-state sample, we developed a coding framework consisting of 16 domains (social gatherings; religious gatherings; funerals; stay at home orders; restaurants; bars; movie theatres; hair salons and barbers; indoor gyms; non-essential retail stores; childcare; K-12 schools; higher education; nursing homes; prisons; voting). These domains were selected to optimize sensitivity and to capture differences in the timing and intensity of social distancing policy responses. While the timing and intensity of social distancing in some of these domains moved in unison, we observed differences among our sample states in imposing social distances between these domains. Policies were coded as either applying statewide or only to a sub-region (e.g., county) of the state, but our analysis was limited to policies adopted by the governor or other statewide executive authority. Therefore, a social distancing policy adopted by a governor that only applies to certain counties was included, but a policy adopted by a mayor was excluded from our analysis.

To optimize sensitivity within domains, we applied an intensity scale comprising 5 levels to each domain. The 5 levels were structured based on the following framework: No Recommendations or Mandates; Recommendations Only; Mandates-Low; Mandates-Medium; Mandates-High; Mandates-Very High. Levels within each domain were defined using domain-specific examples to provide specificity, but also flexibility to allow the scale to evolve over time as social distancing policy responses fluctuate (e.g., by adding masking requirements). We found that domains were sufficiently different that we needed to develop detailed domain-specific intensity scales to improve inter-rater reliability within domains (e.g., different types of restaurant restrictions such as prohibiting on-premises dining but allowing takeaway orders versus allowing on-premises dining but with an occupancy cap). The full social distancing intensity framework, including the detail of domain-specific scales, is included in Exhibit 2 (included at the end of this manuscript). The intensity scale is ordinal, meaning that the numeric relationship between levels of intensity is not necessarily proportional to the effect on social distancing. For example, an intensity scale of 4 in a particular domain does not necessarily mean that the magnitude of social distancing imposed by that level is twice as intense as a score of 2 in the same domain. The numeric ordinal social distancing intensity scale allows for more nuanced quantitative analyses and modeling. We did include a Very High level to allow for an even more stringent level of social distancing that has been seen in other countries (e.g., Lombardy, Italy).

We identified a number of policies adopted by state-level government (e.g., governor Executive Order or Department of Health order) that only applied in a particular county or region within the state. For these days, we calculated a weighted daily average intensity within each domain by 1) coding the number of counties at each intensity level on that day, 2) multiplied the intensity score for those counties by the proportion of counties with that intensity score, and 3) then summed the result.

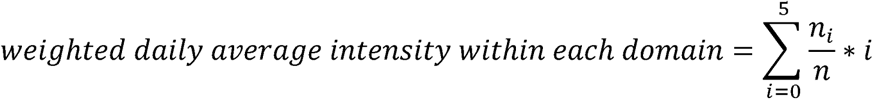

In this equation, i = intensity score, n_i_ = number of counties with that intensity score, and n = total counties in the state. Here, this treats all counties equally, but an alternative option would be to multiply by the proportion of the state’s population living in counties with that intensity score in the second step instead of by the proportion of counties with that score. The overall daily average intensity score is then the mean of the 16 domain-specific scores.

Exhibit 3 provides a summary of key data points from the results of the analysis. As Exhibit 3 illustrates, the states from our sample with the highest average daily intensity were Pennsylvania (4.19), Washington (4.13), Colorado (4.13), California (4.06), and New Jersey (4.06). The states from our sample with the lowest peak average daily intensity were: Georgia (2.94), Florida (3.25), Massachusetts (3.56), and Texas (3.63). The states with the highest daily average intensity on June 19 were: New York (3.47), New Jersey (3.44), Washington (3.15), and California (2.95). The states with the lowest daily intensity on June 19 were: Florida (2.08), Georgia (2.19), Texas (2.50), and Louisiana (2.56).

**Exhibit 2:** Social Distancing Intensity Framework.

| Domain | Level | Score |
| --- | --- | --- |
| Social Gatherings | No recommendations or rules implemented for this domain | 0 |
| Social Gatherings | Recommendations Only | 1 |
| Social Gatherings | Mandates - Low (e.g., prohibits large and very large gathering of more than 100-1000 people) | 2 |
| Social Gatherings | Mandates - Medium (e.g., prohibits medium and large gatherings of more than 10-50 people) | 3 |
| Social Gatherings | Mandates - High (e.g., prohibits small, medium and large gatherings of people not living in the same household) | 4 |
| Social Gatherings | Mandates - Very High (e.g., prohibits all non-essential gatherings outside of household, including gatherings of persons living in the same household) | 5 |
| Religious Gatherings | No recommendations or mandates for this domain | 0 |
| Religious Gatherings | Recommendations only (e.g., minimize large gatherings of more than 250-500 or recommendations to use social distancing + and use remote services where possible) | 1 |
| **Religious Gatherings** | Mandates - Low (e.g., prohibits large religious of more 100-250 people or occupancy caps of approx. 50% or allows church services if social distancing followed) | 2 |
| **Religious Gatherings** | Mandates - Medium (e.g., prohibiting medium and large religious gatherings with more than 10-50 people or occupancy caps of 25% or below) | 3 |
| **Religious Gatherings** | Mandates - High (e.g., prohibits small, medium and large religious gatherings, except for people living in the same household) | 4 |
| **Religious Gatherings** | Mandates - Very High (e.g., no religious gatherings allowed outside of the household) | 5 |
| **Funerals** | No recommendations or mandates for this domain | 0 |
| **Funerals** | Recommendations only (e.g., minimize large gatherings of more than 250/500 or recommendations to use social distancing + hygiene protocols, and use remote services where possible) | 1 |
| **Funerals** | Mandates - Low (e.g., require social distancing or large occupancy cap) | 2 |
| **Funerals** | Mandates - Medium (e.g., prohibits more than 10-50 attendees at funerals; or occupancy caps of approx. 25% or below) | 3 |
| **Funerals** | Mandates - High (e.g., funeral attendees must live in the same household) | 4 |
| **Funerals** | Mandates - Very High (e.g., prohibiting attendees at funerals) | 5 |
| **Stay at Home/Shelter in Place Orders** | No recommendations or mandates for this domain | 0 |
| **Stay at Home/Shelter in Place Orders** | Recommendations Only (e.g., recommendations to stay at home only for vulnerable populations) | 1 |
| **Stay at Home/Shelter in Place Orders** | Mandates - Low (e.g., only applies to limited groups during specified times, strong social distancing mandate but not a stay at home order) | 2 |
| **Stay at Home/Shelter in Place Orders** | Mandates - Medium (e.g., only applies to higher risk groups) | 3 |
| **Stay at Home/Shelter in Place Orders** | Mandates - High (e.g., broadly applicable shelter in place order with exceptions for essential activities) | 4 |
| **Stay at Home/Shelter in Place Orders** | Mandates - Very High (e.g., all residents stay at home, except for essential activities, plus additional measures to reduce social mobility, a curfew, requiring permits when leaving the home, or limiting only 1 person to leave the home at a time) | 5 |
| **Restaurants** | No recommendations or mandates for this domain | 0 |
| **Restaurants** | Recommendations only (e.g., physical distancing in line, limiting hours, limiting occupancy, or closure) | 1 |
| **Restaurants** | Mandates - Low (e.g., require placing tables 6 feet apart but no occupancy cap) | 2 |
| **Restaurants** | Mandates - Medium (e.g., limiting occupancy, limiting hours of operation, limiting on-premise consumption to outdoors only) | 3 |
| **Restaurants** | Mandates - High (e.g., prohibiting on premises dining, but not prohibiting takeout/delivery) | 4 |
| **Restaurants** | Mandates - Very High (e.g., restaurants ordered closed with no takeaway) | 5 |
| **Bars** | No recommendations or mandates for this domain | 0 |
| **Bars** | Recommendations only (e.g., physical distancing in line, limiting hours, limiting occupancy, or closure) | 1 |
| **Bars** | Mandates - Low (e.g., take steps to require physical distancing in lines or plastic screens in front of registers) | 2 |
| **Bars** | Mandates - Medium (e.g., limiting occupancy, limiting hours of operation, limiting on-premises consumption to outdoors only) | 3 |
| **Bars** | Mandates - High (e.g., prohibiting on premises dining, but not prohibiting takeout/delivery) | 4 |
| **Bars** | Mandates - Very High (e.g., bars ordered closed with no takeaway) | 5 |
| **Indoor Movie Theatres** | No recommendations or mandates for this domain | 0 |
| **Indoor Movie Theatres** | Recommendations only (e.g., physical distancing in line, limiting hours, limiting occupancy, or closure) | 1 |
| **Indoor Movie Theatres** | Mandates - Low (e.g., require physical distancing in lines or plastic screens in front of registers) | 2 |
| **Indoor Movie Theatres** | Mandates - Medium (e.g., moderate limits on occupancy such as 50% capacity or only requiring that groups sit 6 feet apart but no cap on occupancy) | 3 |
| **Indoor Movie Theatres** | Mandates - High (e.g., very limited occupancy (e.g., 25% or less)) | 4 |
| **Indoor Movie Theatres** | Mandates - Very High (e.g., must close to the public and prohibit employees from working on premise) | 5 |
| **Hair Salons/Barber Shops** | No recommendations or mandates for this domain | 0 |
| **Hair Salons/Barber Shops** | Recommendations only (e.g., physical distancing in line, limiting hours, limiting occupancy, or closure) | 1 |
| **Hair Salons/Barber Shops** | Mandates - Low (e.g., require physical distancing in lines or plastic screens in front of registers) | 2 |
| **Hair Salons/Barber Shops** | Mandates - Medium (e.g., limiting occupancy or requiring appointments) | 3 |
| **Hair Salons/Barber Shops** | Mandates - High (e.g., closing some premises to the public, but allowing others to continue to operate with other restrictions in place or allowing employees to continue to work onsite to allow for online-based services) | 4 |
| **Hair Salons/Barber Shops** | Mandates - Very High (e.g., closing all premises to the public and prohibiting employees from working on premise) | 5 |
| **Indoor Gyms** | No recommendations or mandates for this domain | 0 |
| **Indoor Gyms** | Recommendations only (e.g., physical distancing in line, limiting hours, limiting occupancy, or closure) | 1 |
| **Indoor Gyms** | Mandates - Low (ex. require physical distancing in lines or plastic screens in front of registers) | 2 |
| **Indoor Gyms** | Mandates - Medium (e.g., limiting occupancy or hours of operation) | 3 |
| **Indoor Gyms** | Mandates - High (e.g., closing some premises to the public, but allowing others to continue to operate with other restrictions in place or allowing employees to continue to work onsite to allow for online-based services) | 4 |
| **Indoor Gyms** | Mandates - Very High (e.g., closing all premises to the public and prohibiting employees from working on premise) | 5 |
| **Non-Essential Retail Stores** | No recommendations or mandates for this domain | 0 |
| **Non-Essential Retail Stores** | Recommendations only (e.g., physical distancing in line, limiting hours, limiting occupancy, or closure) | 1 |
| **Non-Essential Retail Stores** | Mandates - Low (ex. take steps to require physical distancing in lines or plastic screens in front of registers) | 2 |
| **Non-Essential Retail Stores** | Mandates - Medium (e.g., limiting occupancy or hours of operation) | 3 |
| **Non-Essential Retail Stores** | Mandates - High (e.g., allowing curbside pickup or online order processing; only closing some non-essential businesses) | 4 |
| **Non-Essential Retail Stores** | Mandates - Very High (e.g., closing all premises to the public and prohibiting employees from working on premise) | 5 |
| **Childcare/Early Learning** | No recommendations or mandates for this domain | 0 |
| **Childcare/Early Learning** | Recommendations only (e.g., recommend restrictions on visitors and/or screening at entrance) | 1 |
| **Childcare/Early Learning** | Mandates - Low (e.g., restrictions on visitors, screening at entrance; must close buildings temporarily for cleaning if they have a confirmed case) | 2 |
| **Childcare/Early Learning** | Mandates - Medium (e.g.; reductions on class and/or facility capacity to comply with social distancing) | 3 |
| **Childcare/Early Learning** | Mandates - High (e.g., close except for those serving frontline or essential workers; may require special permit) | 4 |
| **Childcare/Early Learning** | Mandates - Very High (e.g., all childcare/early education facilities forced to close, with only very limited exceptions) | 5 |
| **K-12** | No recommendations or mandates for this domain | 0 |
| **K-12** | Recommendations only (e.g., cancel or reschedule large events) | 1 |
| **K-12** | Mandates - Low (e.g., schools required to cancel large events; schools required to close temporarily for cleaning if they have a confirmed case) | 2 |
| **K-12** | Mandates - Medium (e.g., no in-person instruction at public schools, but other services provided at public schools may continue; either no guidance regarding private schools or recommendation that private schools end in-person instruction) | 3 |
| **K-12** | Mandates - High (e.g., no in-person instruction at public or private K-12 schools, but facilities may still be used for other essential services, such as teachers accessing school facilities to facilitate online learning, childcare, distributing pre-packaged lunches) | 4 |
| **K-12** | Mandates - Very High (e.g., no in-person instruction and no admittance to teachers for online learning, but maintenance staff can access for critical facility maintenance) | 5 |
| **Higher Education** | No recommendations or mandates for this domain | 0 |
| **Higher Education** | Recommendations only (e.g., recommendation to cancel or reschedule large events) | 1 |
| **Higher Education** | Mandates - Low (e.g., close buildings temporarily for cleaning if they have a confirmed case; encouraged to examine whether they can continue operating safely; in-person instruction can resume with social distancing) | 2 |
| **Higher Education** | Mandates - Medium (e.g., prohibiting in-person instruction at only public higher education institutions; requiring public higher education institutions to shift to online learning; either no guidance regarding in-person instruction at private higher education institutions or private higher education institutions recommended to end in-person instruction) | 3 |
| **Higher Education** | Mandates - High (e.g., requiring online learning for all public and private higher education institutions, but other services on campus may still continue, such as housing students in dorms, access to teachers to facilitate online learning) | 4 |
| **Higher Education** | Mandates - Very High (e.g., no in-person instruction at private or public higher education institutions; dorms must be vacated; no admittance to teachers for online learning, but maintenance staff can access for critical facility maintenance) | 5 |
| **Nursing Homes** | No recommendations or mandates for this domain | 0 |
| **Nursing Homes** | Recommendations only (e.g., visitor restrictions) | 1 |
| **Nursing Homes** | Mandates - Low (e.g., requiring social distancing in the facility) | 2 |
| **Nursing Homes** | Mandates - Medium (e.g., restriction on number of visitors) | 3 |
| **Nursing Homes** | Mandates - High (e.g., prohibiting non-essential visitors) | 4 |
| **Nursing Homes** | Mandates - Very High (e.g., require full-time "shelter in place" for all facility staff) | 5 |
| **Prisons** | No recommendations or mandates for this domain | 0 |
| **Prisons** | Recommendations only (e.g., physical distancing in facility, visitor restrictions) | 1 |
| **Prisons** | Mandates - Low (e.g., by limiting outside visitors to no more than 3 at a time) | 2 |
| **Prisons** | Mandates - Medium (e.g., only allowing essential visitors, such as attorneys, and banning non-emergency off-site trips); | 3 |
| **Prisons** | Mandates - High (e.g., reduce overcrowding by suspending prisoner transfers; suspending intake of new prisoners from county jails) | 4 |
| **Prisons** | Mandates - Very High (e.g., reduce overcrowding by releasing prisoners who represent low safety risks; prisoners ages 60+ who have release dates within 75 days and whose crimes are nonviolent; commuting sentences for non-violent crimes) | 5 |
| **Voting** | No recommendations or mandates for this domain | 0 |
| **Voting** | Recommendations only (e.g., recommend using existing absentee ballot processes) | 1 |
| **Voting** | Mandate - Low (e.g., extending early voting) | 2 |
| **Voting** | Mandate - Medium (e.g., facilitating mail-in voting by sending out mail-in ballot request forms) | 3 |
| **Voting** | Mandate - High (e.g., requiring that elections be held as mail-in elections, with very limited exceptions) | 4 |
| **Voting** | Mandate - Very High (e.g., postponing elections) | 5 |

**Exhibit 3.** – Social Distancing Intensity by State

| State | Date of First Mandate | Peak Daily Intensity | Date Range of Peak Intensity | No. of Days Between First Mandate and Peak Intensity | No. of Days at Peak Intensity | Intensity on June 19 |
| --- | --- | --- | --- | --- | --- | --- |
| California | 3/11 | 4.06 | 3/24 - 5/7 | 13 | 44 | 2.95 |
| Colorado | 3/12 | 4.13 | 3/27 - 4/26 | 15 | 30 | 2.88 |
| Florida | 3/14 | 3.25 | 4/3 - 5/3 | 20 | 30 | 2.08 |
| Georgia | 3/18 | 2.94 | 4/3 - 4/22 | 16 | 19 | 2.19 |
| Illinois | 3/11 | 3.75 | 3/26 - 4/30 | 15 | 35 | 2.75 |
| Louisiana | 3/12 | 3.69 | 4/9 - 5/14 | 28 | 35 | 2.56 |
| Massachusetts | 3/13 | 3.56 | 3/24 - 5/17 | 11 | 54 | 2.94 |
| New Jersey | 3/13 | 4.06 | 4/1 - 5/12 | 19 | 41 | 3.44 |
| New York | 3/11 | 3.94 | 3/27 - 5/14 | 16 | 48 | 3.47 |
| Pennsylvania | 3/16 | 4.19 | 4/10 - 5/7 | 25 | 27 | 2.72 |
| Texas | 3/13 | 3.63 | 4/13 - 4/30 | 31 | 17 | 2.50 |
| Washington | 3/10 | 4.13 | 4/15 - 5/4 | 36 | 19 | 3.15 |

Exhibit 4 shows the distribution of days at different policy intensity scores within each domain, looking at all states together.

**Exhibit 4.**
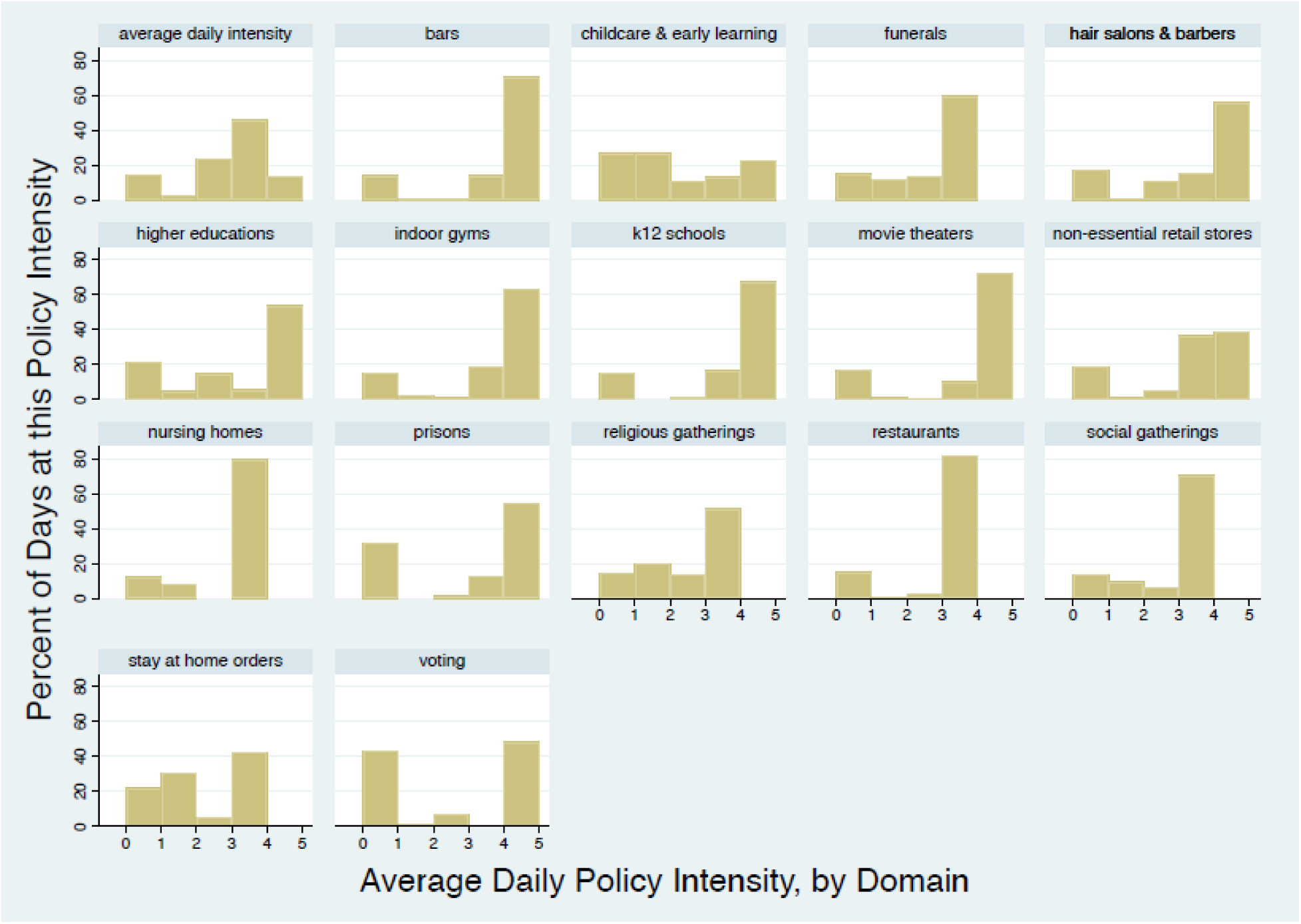
Distribution of Policy Intensity Scores, by Domain

For domains showing sharply bimodal distributions (such as restaurants or movie theaters), the added granularity of this policy coding structure may not add as much informational depth as it does for domains with broader distributions (such as childcare and religious gatherings). However, as states begin to “open back up” we are seeing increased policy specificity even within domains such as restaurants and movie theaters, suggesting that we might expect to see broader distributions across all domains a few months hence.

Exhibit 5 shows the results of the coding displayed for each state in our sample. As illustrated by Exhibit 5 and Exhibit 3, the slope of intensity change during the implementation of social distancing is more severe than during the easing of social distancing restrictions during “re-opening” up through June 19, which is more gradual and reflects the staged re-opening process used by many states.

**Exhibit 5.**
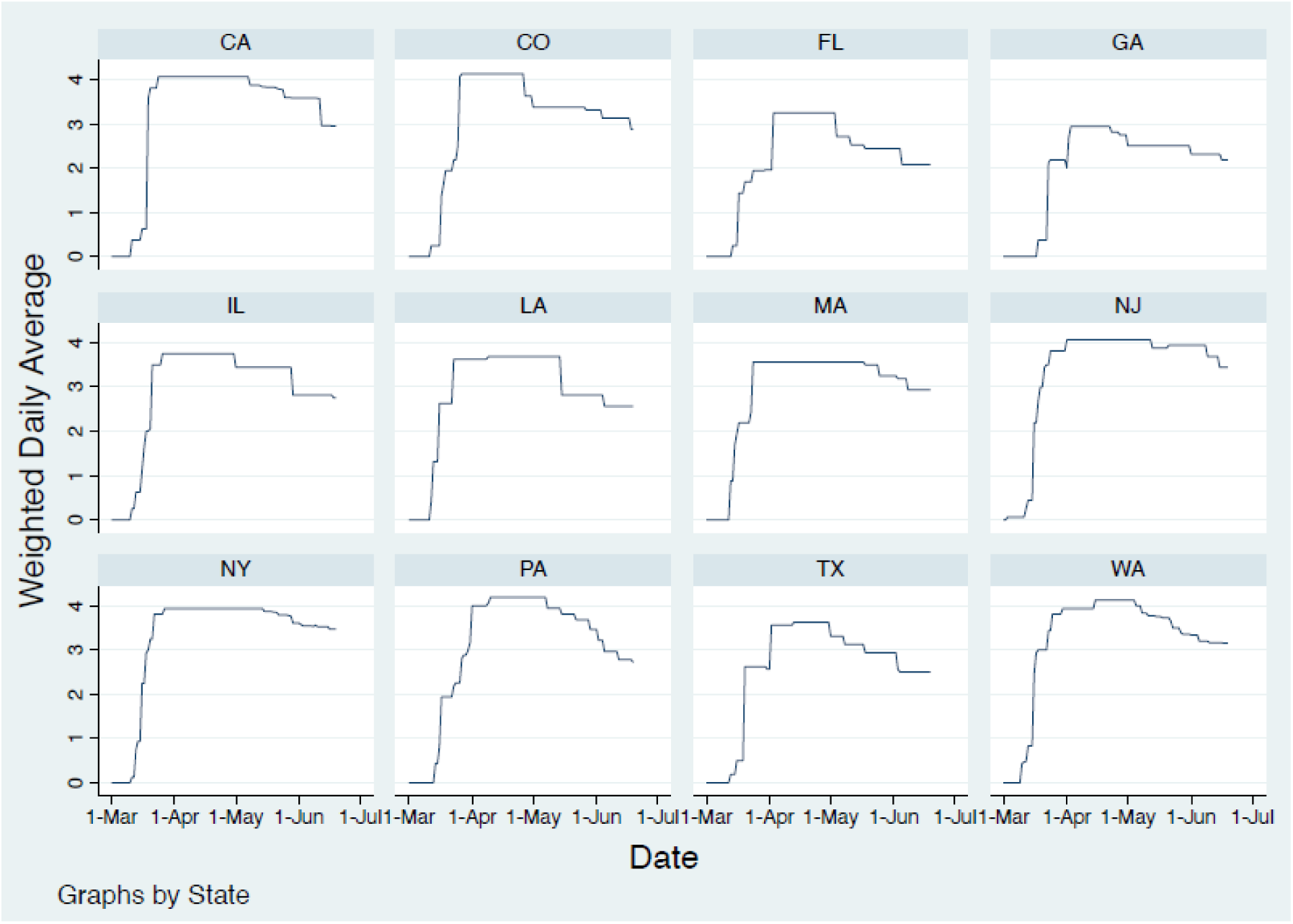
Average Policy Intensity Score, by Date and State

As can be seen in Exhibit 6, we observe a direct temporal relationship between increased policy intensity and a decrease in incidence rates in some states (such as Florida, New York, Washington) but not in others (such as California) – and the potential for a subsequent rise in incidence rates as policy intensity begins to drop back down (Washington, Florida).

**Exhibit 6.**
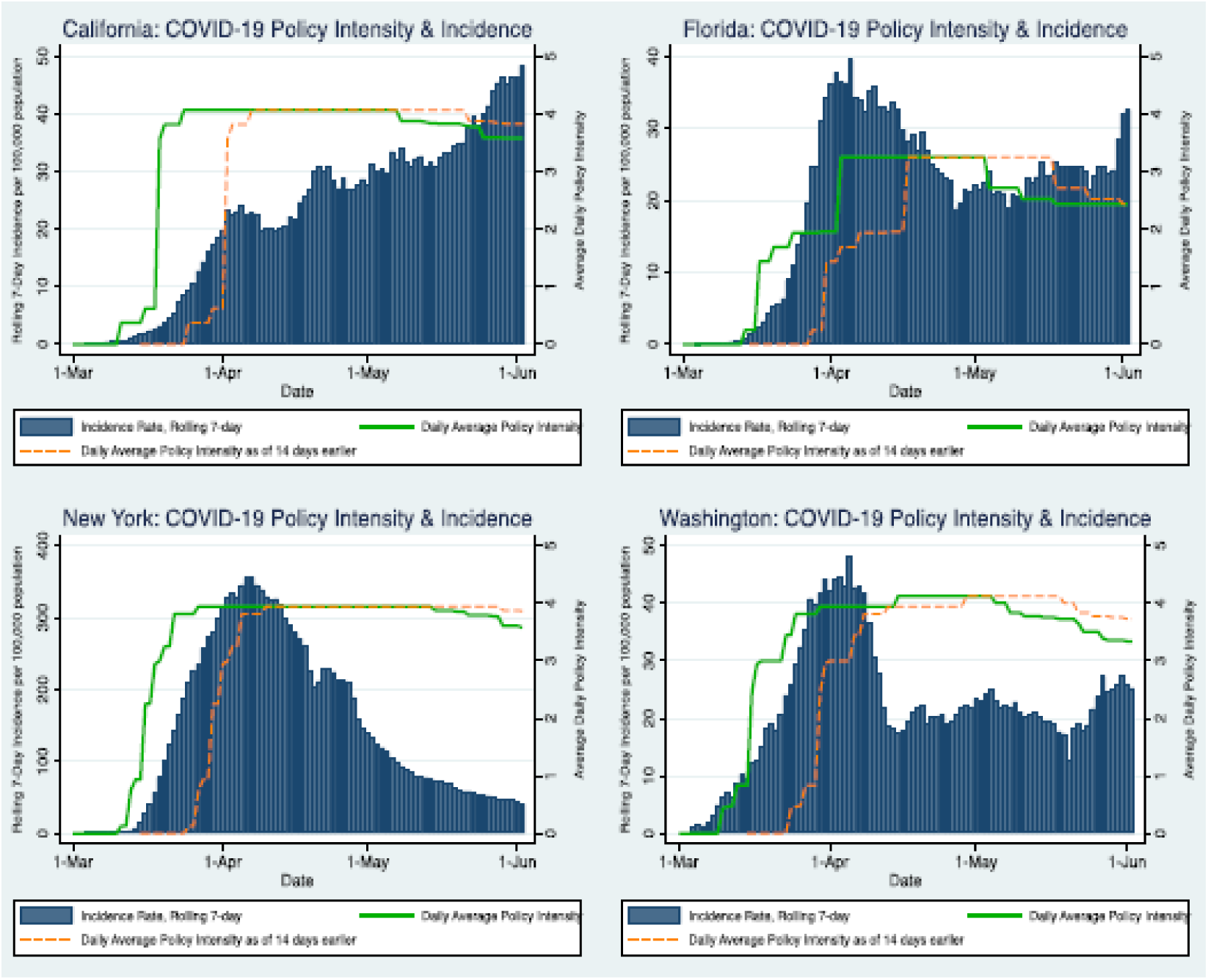
Average Daily Policy Intensity and COVID-19 Incidence Rates: A Four State Comparison

Even in those states which do show drops in COVID-19 incidence rates consonant with the intended policy effects, the relationship appears non-linear, with no decrease in transmission until a policy threshold is met. These graphs also help demonstrate why visualizing the potential lag time in policy impact can be helpful – with the green lines showing the average policy intensity in the state as of the same day as the outcome data, and the orange dashed line showing what the average policy intensity had been 14 days earlier when it had the potential to have an impact on the COVID-19 incidence rate of the current date. As not all policies will have equivalent lag times, it may be useful to visualize a range of different lag times to best identify patterns and relationships.

## Discussion

An effective policy taxonomy should be sufficiently specific and sensitive to facilitate cross-jurisdiction comparisons of policy approaches, monitor material changes to policy frameworks over time, and improve our ability to identify policy variables that improve the relative performance of differing approaches.(17) Taxonomies dig beneath the titles of policies and highlight the underlying and discrete mechanisms of action. The phased re-opening plans of states within our sample illustrate the need for rigorous policy analysis to inform policy decision-making. For example, New York is using a re-opening framework of 4 “Phases,” Pennsylvania is using a Red, Yellow, Green reopening framework, and California is using a series of “Stages.” The relative intensity of each stage/phase/color can vary between states. For example, indoor dining can restart in Washington State in counties reaching Phase 2, but indoor dining did not start in New York regions until they were designated as Stage 3.

The results of the data visualization illustrate the potential advantage of using a policy coding framework that captures nuanced differences in policy approaches. We are also beginning to see similar policy differences in masking policies. Evidence suggests that individual masking may substantially reduce COVID-19 transmission, and states are adopting a range of different types of policies with the goal of increasing individual masking in the population. Some of these policies only recommend wearing masks in public, while other mandate that certain employees wear masks, and still others mandate that members of the public wears masks. Masking policy was outside the scope of our analysis, but the results of our analysis indicate that masking policies, like social distancing policies, should be analyzed in a way that captures the differences in these policy approaches. For example, a tiered masking policy coding framework has been developed by Fullman et al.(12)

A nuanced policy taxonomy will help us more accurately compare the differences between state policy approaches and potentially allow us to isolate approaches that have the most substantial effect, while minimizing social and economic harm. The coding taxonomy presented here provides one approach to capture the nuances of policy responses that may be important for monitoring differences between states as they respond to future outbreak fluctuations.

The policies we analyzed are limited to those adopted by state-level executive authorities (e.g., governors or departments of health); many local jurisdictions have adopted their own measures to institute social distancing sooner or more intensely than required by state authorities. We recorded an increase in the use of county-specific social distancing in a number of our sample states (i.e., Washington, Florida, New York, California, Pennsylvania), especially when they began easing social distancing requirements. For this reason, it may be important to monitor social distancing policy responses at the county-level to have the most accurate view of the current status of social distancing approaches in the United States. Doing so, however, must recognize the interplay between state and local policies and powers (e.g., some state executive orders prohibit more stringent local orders, while other allow them).

We included in our coding framework domains that have not been included in some other social distancing approaches, including domains for voting and prisons. These domains were harder to code using an ordinal scale. Voting presented challenges, because only some states had statewide elections scheduled for March, April, or May. In addition, some states, such as Washington State, already used all-mail in voting prior and unrelated to COVID-19, while others instituted or expanded mail-in balloting as a way to promote social distancing in direct response to the COVID epidemic. Prisons presented other challenges, because it was more difficult to order the relative intensity of some actions (e.g., prohibiting visitors vs. suspending intake from country jails). Nevertheless, the authors believe these are important domains to analyze for comparative analysis purposes, and we have included them in this proposed scale. An alternative approach for this domain could be to use a normal scale for some of these domains (e.g., 1 point if a sub-domain action is present), which would not require assigning a relative order of intensity, but would still provide a more nuanced picture of the response within that domain. Another result of having a larger number of domains is that some domains were subject to social distancing together or indirectly by some state policies, while other states had more specific policy guidance that specifically identified different types of locations. For example, in some states, social distancing at commercial businesses was restricted through a ban on gatherings of certain size or stay at home order (e.g., California), while others ordered specific types of businesses to close in addition to issuing a stay at home order (e.g., Louisiana).

In addition, we did not assess the extent of implementation or enforcement of these policies. Implementation of public policies often varies from the written word for a variety of reasons, including limited enforcement resources or local interpretation. This may be especially true for some social distancing policies, such as bans on religious services, which rely on broad public compliance with very little enforcement by government officials. For this reason, cultural views of the importance of social distancing are important mediators for implementing the social distancing policies we analyzed. The effectiveness of social distancing policies may also change over time as people accommodate or, alternatively, grow tired of the restrictions.

Policy makers have increasingly relied on disease modeling that account for policy decisions to inform their COVID-19-related decisions. For example, Dr. Deborah Birx, the Coronavirus Response Coordinator, has repeatedly cited the COVID-19 projections prepared by the University of Washington’s Institute for Health Metrics and Evaluation (IHME) at White House COVID Task Force briefings.(18) The methods used by various modeling efforts for linking COVID policies to projected outcomes (e.g., rates of infection, hospitalization, or death), however, have been quite divergent. The policy coding framework we developed has not yet been used to populate disease modeling projections, and it is possible that the increased descriptive power of this framework does not yield more accurate disease models. Additional analyses are underway to test whether this coding approach provides greater power to quantitative analyses.

COVID-19 presents an opportunity to develop standardized approaches for policy taxonomies and coding processes to facilitate more rigorous analysis and improve disease modeling efforts that rely on sensitive prognostic variables, such as assumptions about future government policy responses. Based on our experience developing this coding framework, we make the following recommendations about policy coding methods to improve comparative policy analyses and disease modeling:

- Develop a taxonomy that captures the nuances and gradients between policy approaches (domains, sub-domains) that may have a material effect on the effectiveness of such policies;
- Consider an intensity or stringency ordinal scale when appropriate to facilitate comparison between jurisdictions but recognize that not all domains are necessarily appropriate for an ordinal scale – if an ordinal scale is not appropriate, consider a normal scale, which does not have an ordering;
- Clearly define scales and domains to improve inter-rater reliability and to facilitate updates as policy approaches evolve;
- Identify specific policies relied upon (e.g., title, date) and make copies of those policies available on a website to allow secondary review and broader access to the primary source policies;
- Include interpretation notes so reviewers and other researchers are aware of how ambiguities were resolved or interpreted
- Capture PDF and/or screenshots of policies when conducting reviews to ensure access even after websites have been updated or revisions (especially for rapidly evolving policy responses, such as the responses to COVID-19);
- Encourage public review of coding and interpretation to help validate policy interpretations and coding;
- Consider inviting co-investigators from jurisdictions involved in comparative analysis to validate policy interpretations;
- Acknowledge that implementation or enforcement of policies may vary widely across policy domains and jurisdictions; and
- Consider describing the governance structure in the jurisdiction being coded to facilitate understanding of the decision-making authority of state versus local jurisdictions and among state agencies (e.g., the authority of the governor versus the state health officer or education secretary).

## Conclusion

COVID-19 has presented once of the most rapid and intense phases of public health policy development and implementation in the last century. Often these policy decisions have had to be made with imperfect evidence under very tight timelines. As we continue to live with COVID-19, attempt to ease social distancing, and allow greater economic activity and social interaction, accurate disease models and policy monitoring systems will be critical to evidence-based policymaking. The effectiveness of state-level policies calling for social distancing may be influenced by a variety of factors, including public leadership, local/county policy responses, and local culture. Nevertheless, state-level policy decision-making is an important part of the story of social distancing in response to COVID-19. We hope this detailed social distancing intensity framework and the associated recommendations will provide a more granular view of social distancing approaches and contribute to improving the governmental response to COVID-19.

## Data Availability

The data referred to in the manuscript is available at the indicated website.

https://openicpsr.org/openicpsr/project/120598/version/V1/view

## Exhibit List

EXHIBIT 1: (Table)
Caption: Existing COVID-19 Policy Responses Coding Frameworks Source/Notes: Publicly available literature.
EXHIBIT 2 (table)
Caption: Social Distancing Intensity Framework
Source/Notes: Primary source policies posted to state government websites EXHIBIT 3 (table)
Caption: Social Distancing Intensity By State
Source/Notes: Authors’ analysis of state policies posted to government websites.
EXHIBIT 4 (figure)
Caption: Distribution of Policy Intensity Scores, by Domain
Source/Notes: Authors’ analysis of state policies posted to government websites.
EXHIBIT 5 (figure)
Caption: Average Policy Intensity Score, by Date and State
Source/Notes: Authors’ analysis of state policies posted to government websites.
EXHIBIT 6 (figure)
Caption: Average Daily Policy Intensity and COVID-19 Incidence Rates: A Four State Comparison
Source/Notes: Authors’ analysis of state policies posted to government websites.

